# Predictors of longitudinal cognitive ageing from age 70 to 82 including *APOE* e4 status, early-life and lifestyle factors

**DOI:** 10.1101/2022.02.25.22271448

**Authors:** Janie Corley, Federica Conte, Sarah E. Harris, Adele M. Taylor, Paul Redmond, Tom C. Russ, Ian J. Deary, Simon R. Cox

**Affiliations:** Lothian Birth Cohorts, Department of Psychology, University of Edinburgh, UK; Department of Psychology, University of Milano-Bicocca, Milan, IT; Alzheimer Scotland Dementia Research Centre, University of Edinburgh, UK

## Abstract

Discovering why some people’s cognitive abilities decline more than others is a key challenge for cognitive ageing research. The most effective strategy may be to address multiple risk factors from across the life-course simultaneously in relation to robust longitudinal cognitive data. We conducted a 12-year follow-up of 1091 (at age 70) men and women from the longitudinal Lothian Birth Cohort 1936 study. Comprehensive repeated cognitive measures of visuospatial ability, processing speed, memory, verbal ability, and a general cognitive factor, were collected over five assessments (age 70, 73, 76, 79, and 82 years) and analysed using multivariate latent growth curve modelling. Fifteen life-course variables were used to predict variation in cognitive ability levels at age 70 and cognitive slopes from age 70 to 82. Only *APOE* e4 carrier status was found to be reliably informative of general- and domain-specific cognitive decline, despite there being many life-course correlates of cognitive level at age 70. *APOE* e4 carriers had significantly steeper slopes across all three fluid cognitive domains compared with non-carriers, especially for memory (β = −0.234, P< 0.001) and general cognitive function (β = −0.246, P<0.001), denoting a widening gap in cognitive functioning with increasing age. Our findings suggest that when many other candidate predictors of cognitive ageing slope are entered en masse, their unique contributions account for relatively small proportions of variance, beyond variation in *APOE* e4 status. We conclude that *APOE* e4 status is important for identifying those at greater risk for accelerated cognitive ageing, even among ostensibly healthy individuals.

## Introduction

With advancing age, a pattern of decline is observed across a multitude of cognitive domains, the magnitude differs across domains, and individual differences in rate of cognitive change are substantial [1-2]. Some cognitive abilities, such as vocabulary, remain relatively intact into later life. Other, complex cognitive processes such as processing speed, reasoning, and memory—which require the manipulating of mental data—begin to decline from early adulthood [3-5], and some of these changes are underpinned by a general factor of cognitive ageing [6-8]. Deterioration in cognitive abilities is linked to impairments in older adults’ everyday functions [9], quality of life [10], and health [11], as well as spiralling healthcare costs. Better understanding of long-term cognitive trajectories and their determinants, could inform public policy regarding targeted interventions for those adults at greatest risk of rapid decline, and of progression to Alzheimer’s Disease and other dementias [12], as well as protective factors for staying sharp in later life.

The determinants of individual differences in age-related cognitive decline are likely to include genetic and early-life factors, adult socio-economic status, and health [13-15], though estimates differ with respect to their individual contributions. Risk of accelerated cognitive decline increases with age, cerebrovascular disease, cardiovascular risk factors (e.g. diabetes, obesity) and heart disease [16], but these factors only partially account for cognitive decline risk among the general population [14]. The *APOE* (apolipoprotein) e4 allele is a well-established genetic risk factor for Alzheimer’s disease [17-18], however, the reported effects of *APOE* e4 across the full spectrum of cognitive functioning are highly inconsistent and there is disagreement about whether or not *APOE* e4 influences the rate of cognitive decline in healthy adults [19-25]. Despite a broad corpus of research literature on the role of behavioural risk factors in mitigating age-related cognitive decline, such as smoking, physical activity, alcohol, and diet [3, 26-27], the evidence is patchy and often classed as low to moderate quality [10]. Importantly, many of the effect sizes are small, some are poorly replicated, and findings are often partly, or wholly, attributed to reverse causation, where prior cognitive ability causes variation in the supposed cause of cognitive ability in later adult life [13].

Cognitive decline trajectories are likely to be the result of an accumulation of small effects from numerous individual genetic and environmental risk factors across the life-course [28]. Even smoking, for which there is consistent and demonstrable evidence of an adverse effect on cognitive and brain ageing [29-31], generally accounts for around only 1% of the variance in cognitive decline, similar in magnitude to the estimated effect size of *APOE* e4 on cognitive change from childhood to adulthood [32]. Given that many risk factors for cognitive decline are correlated [33], modelling these potential predictors together, i.e. simultaneously, may be a more useful approach than focussing on single candidate determinants (such as one individual lifestyle or health factor). Multivariate modelling acknowledges the multicollinearity among risk factors and provides more insight into their relative contributions to cognitive change. The very few studies to have tested multiple risk factor models of longitudinal (multi-domain) cognitive decline report few consistent correlates of cognitive change across abilities [34-35]. In the same sample as in the current study—the Lothian Birth Cohort 1936—an earlier multivariate analysis by Ritchie et al. showed that faster rates of decline from age 70 to 76 years were observed in *APOE* e4 carriers, men, and those with poorer physical fitness for some, but not all, cognitive domains [36]. The present study doubles the time frame of that paper (from six to twelve years of follow up), using data from five sampling occasions over a more critical period for accelerated cognitive decline and dementia [37-38], and includes several additional potential predictors (depression, living alone, physical activity, stroke). Having previously identified *APOE* e4 status as an independent predictor of cognitive change in this cohort and elsewhere, we also perform separate trajectory analyses by *APOE* e4 status.

A major challenge in understanding the predictors of cognitive ageing trajectories is the difficulty in disentangling actual cognitive change from lifelong levels of performance (which are conflated in cross-sectional data) and partitioning the variance appropriately [8]. Longitudinal studies with years of repeated cognitive measures are key to understanding the dynamics of cognitive change as people age and can thereby suggest how outcomes might be linked to putative influences. Studies with longer sampling periods and multiple observations increase the power to detect reliable effects and provide more robust evidence than those with few measurement points [15]. Good characterisation of cognitive abilities is crucial, using several cognitive tests for each domain which are sensitive to subtle, age-related, cognitive changes. The accurate estimation of slope trajectories also requires appropriate statistical methods which take into account the complexity of longitudinal, observational data.

Here, we apply multivariate latent growth curve modelling to comprehensive cognitive data collected at five time-points over 12 years to characterise trajectories of cognitive change from ages 70 to 82, and their determinants, in a sample of community-dwelling older adults living in Scotland—the Lothian Birth Cohort 1936 (LBC1936)—for whom there are cognitive function scores from early life. Studies that can account for early-life cognitive ability are rare and valuable with respect to the temporal primacy of cognitive changes (see **Box 1** for other advantages of the study design). Trajectories of cognitive function were evaluated for four major domains of cognitive ability—visuospatial ability, processing speed, and memory (characterising fluid intelligence), and verbal ability (characterising crystallized intelligence). A wide range of potential predictors of cognitive decline from the previous literature were selected covering several categories: early-life (education, childhood IQ); demographic (age, sex, living alone, socio-economic status); lifestyle (smoking, physical activity, body mass index, alcohol), health (cardiovascular disease, diabetes, stroke); depressive symptoms; and *APOE* e4 carrier status. We also examine associations between predictors and a general factor of cognitive function which accounts for the shared variance across the cognitive domains.

## Materials and Methods

### Participants

The Lothian Birth Cohort 1936 (LBC1936) [39-41] is a community-dwelling sample of 1091 men and women in Scotland, being studied in later life for the purposes of assessing the nature and determinants of cognitive and brain ageing, for whom childhood IQ scores are available. Participants were recruited into the LBC1936 in 2004-2007 at the age of ∼70 years, and have so far been followed-up every three years at ages 73 (N = 866), 76 (N = 697), 79 (N = 550), and 82 (N = 431). Socio-demographic, medical history, physical function, blood-derived biomarkers, cognitive function, and lifestyle data were collected at all five waves of in-person testing. For the purposes of the current study, ‘completers’ refer to participants who remained in the study at the age 82 assessment (N = 431), and non-completers (N = 660) were those who dropped out or died at any point between baseline testing and age 82 follow-up. The LBC1936 are surviving participants of the Scottish Mental Survey of 1947 (SMS1947), which tested the mental ability of 70,805 11-year old children born in 1936, using a general intelligence test (The Moray House Test (MHT)) [42]. MHT scores were recorded and archived by the Scottish Council for Research in Education (SCRE), and were made available to the LBC1936 study. For the current study, the MHT score from age 11 was age corrected and converted into a standard IQ-type score for the sample (mean = 100, SD = 15)—henceforth referred to as age 11 IQ—and used a measure of childhood cognitive ability.

#### Ethical approval

Ethical approval was obtained from the Multicentre Research Ethics Committee for Scotland (baseline, MREC/01/0/56), the Lothian Research Ethics Committee (age 70, LREC/2003/2/29), and the Scotland A Research Ethics Committee (ages 73, 76, 79, 82, 07/MRE00/58). All participants provided written informed consent.

### Cognitive Measures

Cognitive function was measured using a detailed battery of well-validated cognitive tests administered at age 70 (baseline), and repeated at ages 73, 76, 79, and 82 years, by trained psychologists [39]. Most of the cognitive tests derive from the Wechsler Adult Intelligence Scale III-UK edition [43] and the Wechsler Memory Scale III-UK edition (WMS-IIIUK) [44]. According to previous work examining their correlational structure [7], the cognitive tests were categorised into four domains of cognitive functioning. Visuospatial ability was measured using Block Design and Matrix Reasoning (WAIS-IIIUK) and Spatial Span (Forwards and Backward) (WMS-IIIUK). Processing Speed was measured using Digit-symbol Coding and Symbol Search (WAISIII-UK) and two experimental tasks: Choice Reaction Time [45]; and Inspection Time [46]. Memory was measured using Verbal Paired Associates and Logical Memory (WMSIII-UK) and Digit-span Backwards (WAIS-IIIUK). Verbal ability was measured using the National Adult Reading Test [47], the Wechsler Test of Adult Reading [48], and Verbal Fluency [49]. A general cognitive factor was constructed based on the shared variance between the four cognitive domains (see Statistical Analysis). The Mini-Mental State Examination (MMSE) [50], widely used as a screening test for possible dementia, was administered at each wave of testing.

### Predictor Measures

Potential risk or protective factors for cognitive decline in later life were identified following a review of previous analyses of the cohort and other population studies; values were obtained from participants’ baseline assessment at age 70.

#### Demographics and early-life

These predictors included age (in days), sex, age 11 IQ score (described above), education (years of formal full-time schooling), living alone (yes/no), and socioeconomic status (SES). SES was coded into six categories based on participants’ highest achieved occupation: 1 (highest professional occupations) to 5 (unskilled occupations), with 3 (skilled occupations) divided into 3N (non-manual) and 3M (manual), using the Classification of Occupations, 1980 [51].

#### Lifestyle

Smoking was coded as current, former or never smoker. Physical activity was coded according to six categories: 1 (‘moving only in accordance with household chores’; lowest level of activity) to 6 (‘keep fit or aerobic exercise several times a week’; highest level of activity). Alcohol units per week were calculated using data collected at interview. Body mass index was calculated using height and weight measurements taken by trained nurses at the time of assessment. Depressive symptoms were measured using the Depression subscale of the Hospital Anxiety and Depression Scale [52] with a score range of 0-21.

#### APOE e4 and health indicators

*APOE* e4 carrier status (yes/no) was determined by genotyping at two polymorphic sites (rs7412 and rs429358) using TaqMan technology. Health indicators included self-reported history (yes/no) of cardiovascular disease (CVD), diabetes, and stroke.

### Statistical Analysis

#### Descriptive

Descriptive statistics are presented for the full sample, and ANOVA and Chi-square tests were used to identify differences in baseline characteristics between groups (completers vs non-completers and deaths vs non-deaths).

#### Trajectories of cognitive decline

We applied latent growth curve (LGC) modelling to the data to investigate level (i.e. intercept, age 70) and trajectories of change (i.e. slope, age 70 to 82) in cognitive functioning across all five waves of testing. A SEM-based ‘factor-of-curves’ [53] approach was used, as has been done previously in this cohort [36, 54] which postulates the existence of common latent variables of cognitive change that underlie the distribution of explicit or observable variables (individual cognitive tests). In our models, we used the average time lag (in years) between the waves: (0, 2.98, 6.75, 9.81, 12.53) as the path weights for calculation of the slope factor. The path from the slope factor to baseline test score was set to zero.^1^ LGC analyses were conducted using the latent variable analysis package ‘lavaan’ [55] in R version 4.0.2 (R Foundation for Statistical Computing, Vienna, Austria) and the code is available online (https://www.ed.ac.uk/lothian-birth-cohorts/discoveries/summary-data-resources). First, we fitted a single parallel process growth curve model at the level of the thirteen individual cognitive tests; intercepts and slopes were correlated, but no hierarchical factor structure was imposed. Second, we fit separate growth curve models for each cognitive domain: visuospatial ability; processing speed; memory; and verbal ability. Here, the latent intercepts and slopes of each cognitive test load onto superordinate latent intercepts and latent slopes of their respective cognitive domains. The cognitive domain models were run for the full sample and also by *APOE* e4 carrier status (yes/no). Unstandardized (beta) estimates, standard errors, p-values, and standard deviation (SD) change per year, are reported.

#### Predictors of cognitive level and slope

We fit both univariate and multivariate risk factor models to the cognitive data to address which factors might contribute to individual differences in cognitive level (age 70) and slope (age 70 to 82). First, univariate LGC models were fit to test the associations of the fifteen individual life-course predictors (alongside age and sex) with each cognitive domain, i.e. without the other variables present in the model. For our main analyses, we fit multivariable LGC models to investigate the relative contributions of each risk factor to cognitive level and slope for each cognitive domain. By including all of the predictors simultaneously, we were able to compare the degree of variance in cognitive level and change accounted for by each risk factor, whilst controlling for the effects of all the other predictors in the model. We ran an additional model representing a general cognitive factor; this hierarchical model was fitted using the latent intercepts and slopes of each of the four cognitive sub-domains, and represents the shared (common) variance between them (**Figure 1** illustrates the hierarchical model framework for general cognitive function). Fully standardised estimates, obtained using the ‘standardizedSolution’ function in lavaan, are presented.

**Figure 1.**
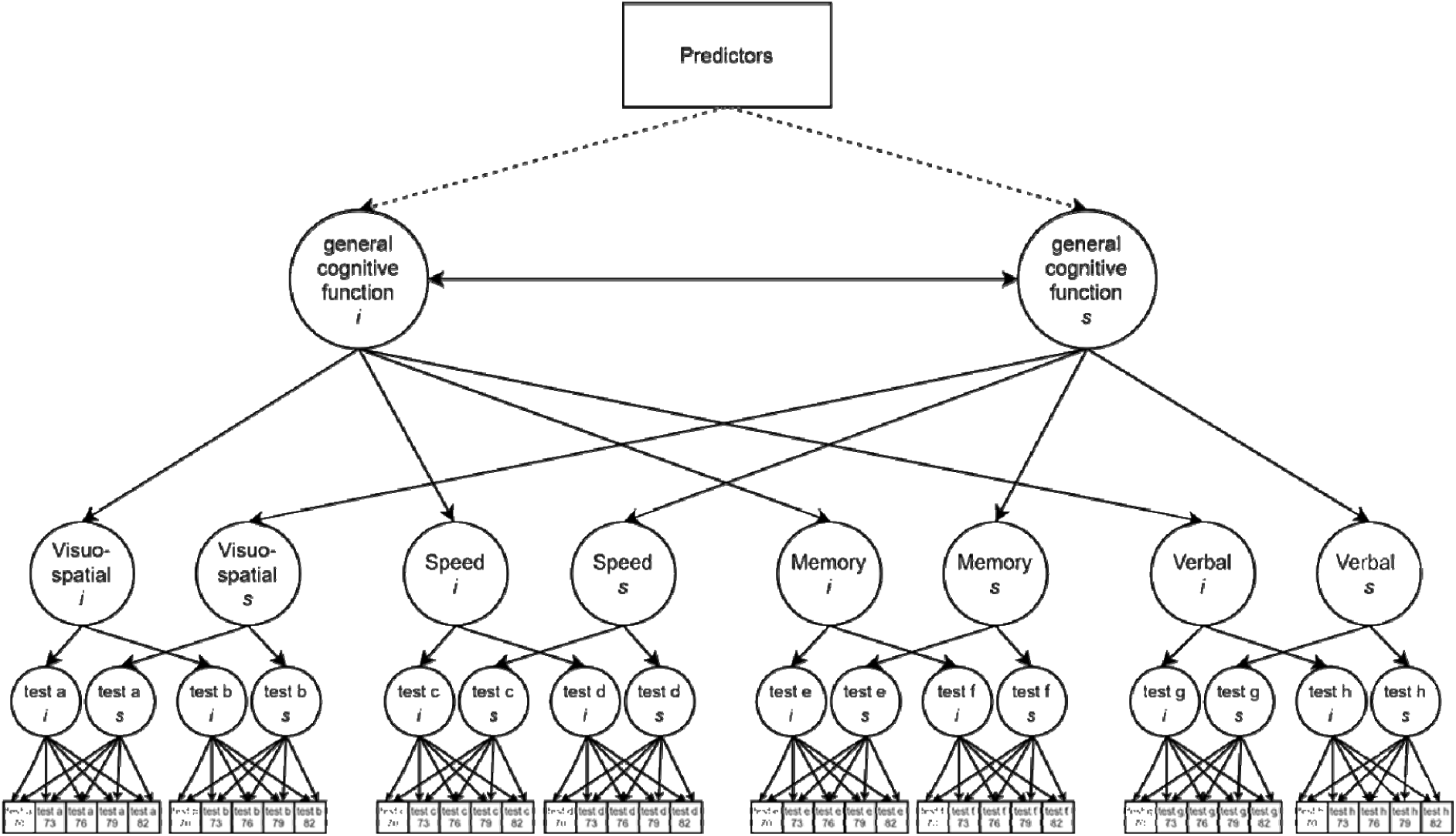
Schematic latent growth curve model in which predictors are associated with the intercept and slope of a latent factor of general cognitive function. A latent growth curve was estimated across five waves of data in a hierarchical model based on the intercepts and slopes of four cognitive domains. For illustrative purposes, not all tests are shown. The regressions of predictors (represented by the dotted lines) on general cognitive function intercept (*i*) and slope (*s*) were the associations of interest. Note that separate models were run for each of the four cognitive domains where the regressions on ‘domain’ intercept (*i*) and slope (*s*) were the associations of interest.

#### Gaussian confounds analysis

With a large set of predictors, as in the current study, we increase the proportion of variance that can be explained in our cognitive outcomes by chance. In order to test whether or not the variance accounted for by the real predictors was comparable to a set of random predictors, we generated a set of Gaussian noise (and random binary) variables and entered them into the LGC models in place of the real predictors, and compared the model R^2^ for each domain. To optimise comparability, we ensured that the same number of continuous vs binary variables were used, and that the patterns of missingness were matched with the real-world predictors.

#### Sensitivity analyses

We repeated the same baseline prediction models in three sensitivity analyses excluding: 1) individuals who reported a subsequent-to-baseline diagnosis of dementia (all participants were dementia-free at baseline); 2) individuals with an MMSE score <24 at any wave, as an indicator of possible pathological ageing; 3) deaths to follow-up (using linkage data obtained via National Health Service Central Register up to April 2021, provided by the National Records of Scotland).

#### Model fit and significance statistics

Models were run using full information maximum likelihood (FIML) estimation to ensure models used all available data to partially mitigate the bias of estimated trajectories and associations by participation bias. Instances of non-significant negative residual variance were set to 0 to allow models to converge upon within-bounds estimates. Model fit was tested using three indices of absolute fit: Comparative Fit Index (CFI) and Tucker-Lewis Index (TLI) (values > 0.95 considered acceptable); and Root Mean Square Error of Approximation (RMSEA) (values <0.06 considered acceptable). Correction for multiple testing was applied across LGC prediction models using the False Discovery Rate (FDR) [56] adjustment, and results marked in bold type are FDR-significant.

## Results

### Descriptive

Baseline characteristics and cognitive test scores for the sample (N = 1091) are shown in **Table 1**. Baseline age was 70 years (mean = 69.5, SD = 0.8), 49.8% of the sample were women, and mean number of years of education was 10.7 (SD = 1.1). *APOE* e4 allele carriers (N = 306) made up 28.0% of the overall sample. *APOE* e4 data were missing for 63 participants (5.8% of the sample). Characteristics are also presented according to completer status, and mortality status by the end of the follow-up period. Participants with fewer follow-up examinations (i.e. non-completers, N = 660) had less education, lower childhood IQ, lower SES, lower physical activity, higher BMI, more depressive symptoms, and were more likely to be a smoker, have a history of CVD, diabetes, and stroke. Non-completers had significantly lower cognitive test scores at baseline than completers. Participants lost to follow-up as a result of death (N = 403) had a lower age 11 IQ, lower SES, lower physical activity, higher BMI, higher alcohol intake, more depressive symptoms, and were more likely to be male, a smoker and to have a medical history of CVD, diabetes and stroke, than those who survived to follow-up. Mean cognitive test scores at baseline were significantly lower in those who had died, compared with the survivors, except for Verbal Pairs (a memory test) and Verbal Fluency (a verbal ability test), for which the group differences were not significant. As noted above, we used FIML estimation in our LGC analyses to reduce any bias due to missingness.

**Table 1.**
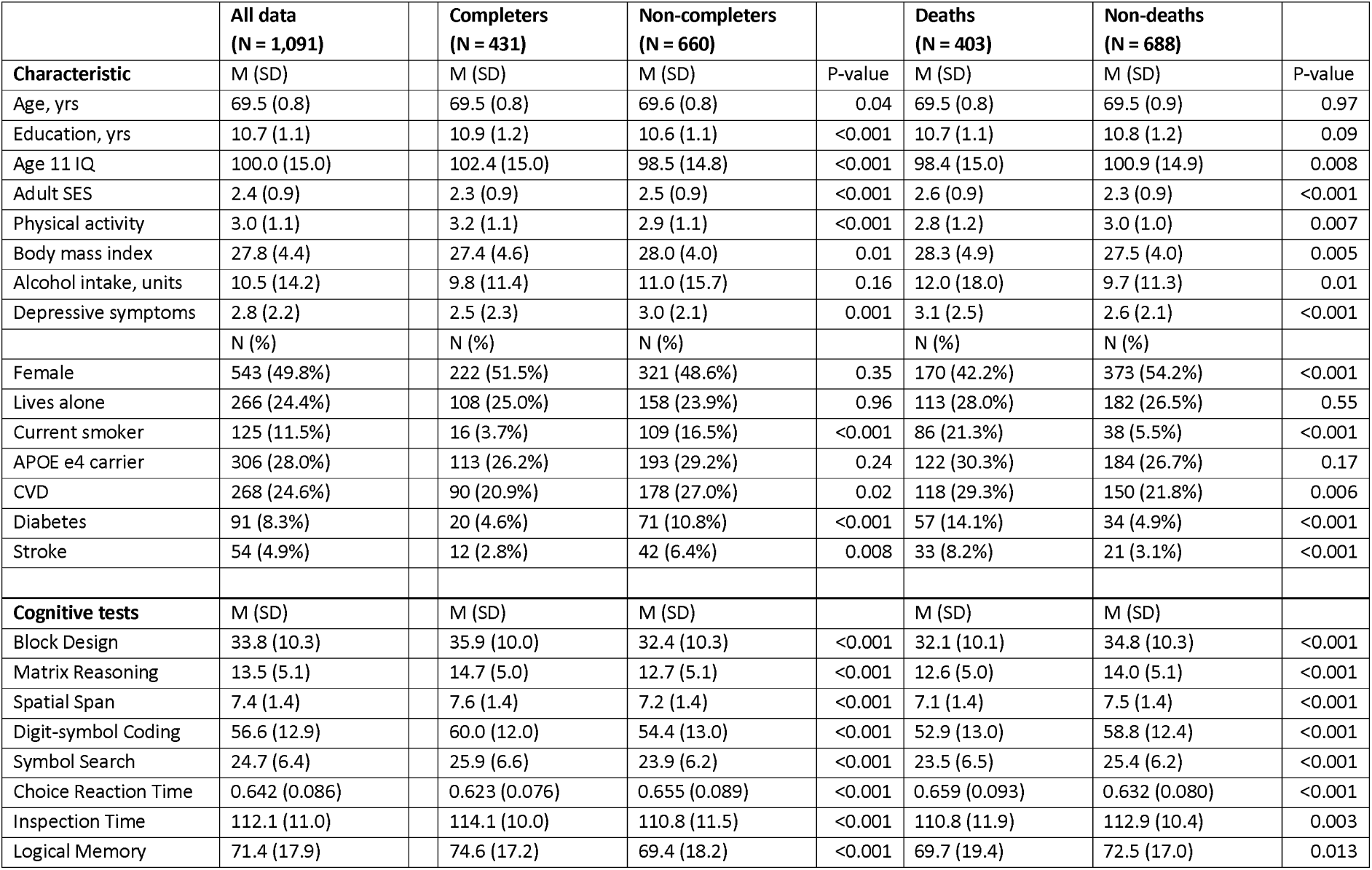
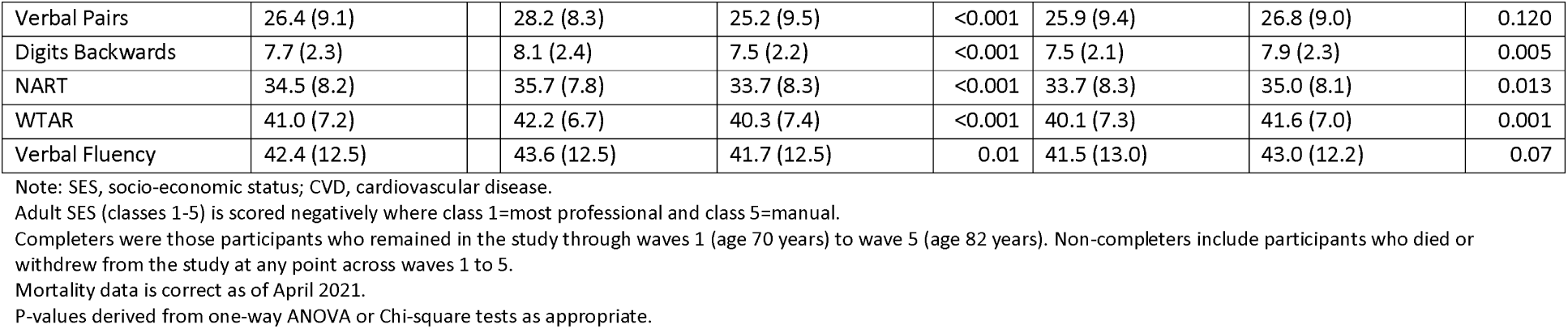
Baseline characteristics of participants overall, and according to completer status and mortality status at the end of follow-up: the Lothian Birth Cohort 1936

A summary of the longitudinal cognitive test scores for the whole sample is presented in **Table 2**. Mean cognitive test scores declined between age 70-baseline and age 82 follow-up, except for two memory tests (Logical Memory and Verbal Pairs) and the verbal ability tests (NART, WTAR, and Verbal Fluency), which were marginally higher at age 82. Logical Memory and Verbal Pairs contain memorable material, which may have resulted in a rise in score in at least the second occasion of testing as a result of practice effects. All three verbal ability tests showed little change over time, and small increases in mean scores at age 82 compared with baseline. Further descriptive information about the cognitive tests scores for completers only, and by *APOE* e4 carrier status, is provided in the Supplementary materials. In the subset of completers only (**Supplementary Table S1**; this has the advantage that the same individuals appear at all waves), all of the mean cognitive test scores were lower at age 82 follow-up compared with baseline with the exception of WTAR (where the mean score was the same), and NART and Verbal Fluency which were slightly higher at follow-up. Note that Choice Reaction Time is scored negatively, such that a higher score indicates a slower reaction time. Mean cognitive test scores at age 70 and age 82 differed according to *APOE* e4 carrier status (**Supplementary Table S2**). At age 70, *APOE* e4 carriers had significantly lower scores on Matrix Reasoning, Spatial Span and Inspection Time than non-carriers. By age 82, *APOE* e4 carriers had significantly lower scores on Block Design, Matrix Reasoning, Spatial Span, Digit-symbol Coding, Symbol Search, Choice Reaction Time, Logical Memory, Verbal Pairs, and Digits Backwards, and the differences were larger in magnitude than at age 70. **Figure 2** plots the linear fitted regression lines through the raw test data for each of the cognitive tests by *APOE* e4 carrier status (non-linear fitted lines through the same data can be found in **Supplementary Figure 1**).

**Table 2.**
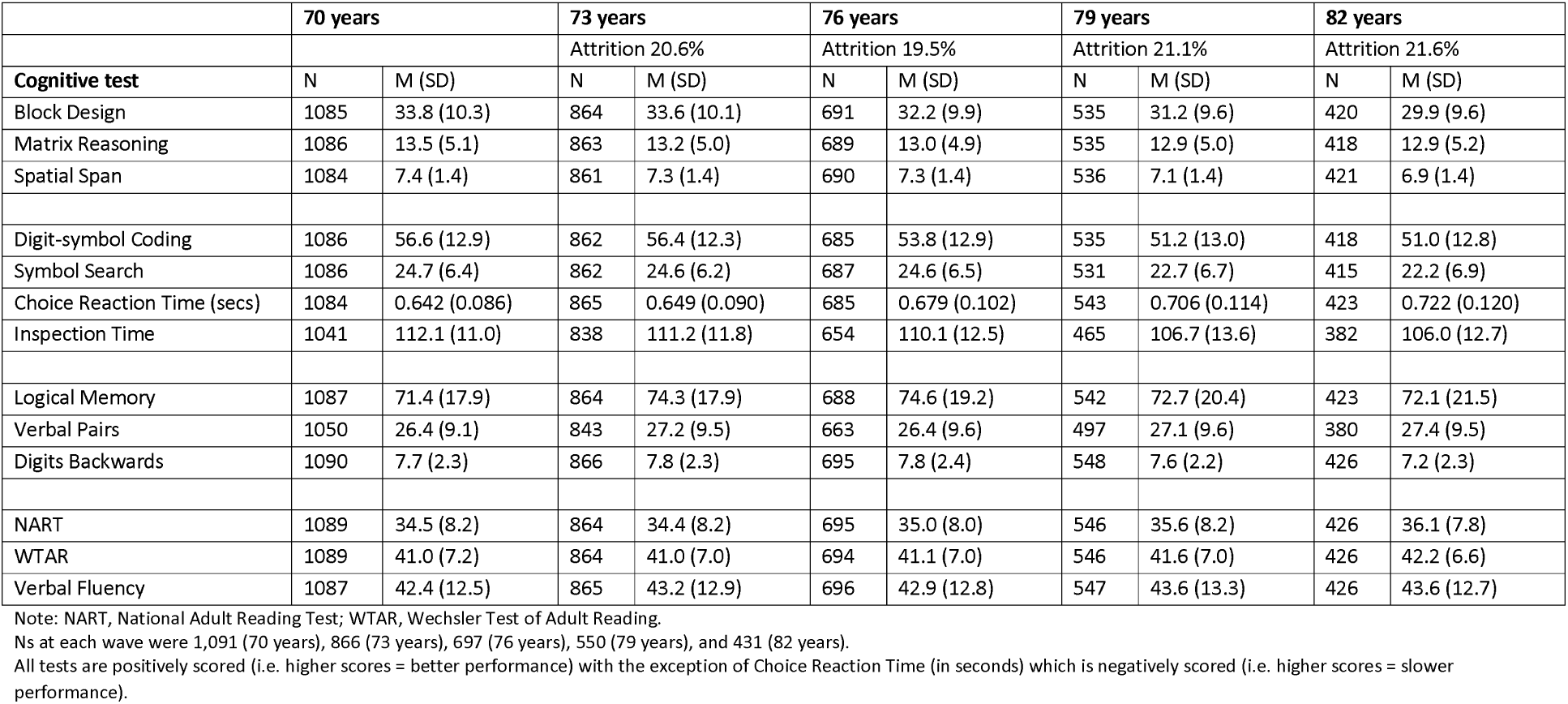
Longitudinal cognitive test scores for all participants

**Figure 2.**
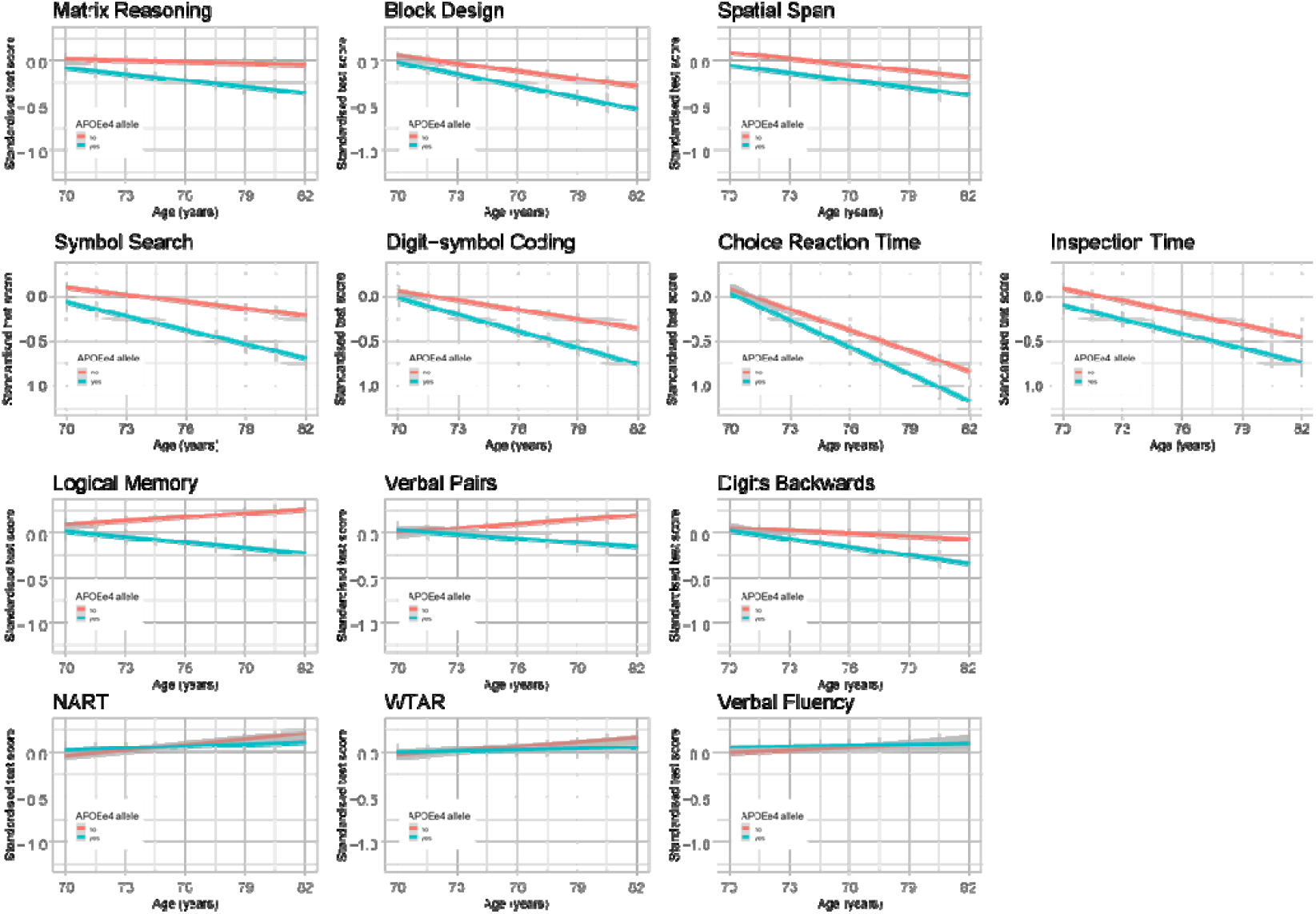
Plots of the regression lines fitted through the raw data, normalised for baseline score, to illustrate the differences in trajectories of cognitive change with age by APOEe4 carrier status (with shaded 95% confidence intervals). Red = non-carrier, blue = carrier.

### Trajectories of cognitive decline

#### Individual cognitive test scores

First, we simultaneously tested whether there was significant ageing-related mean change in each of the thirteen individual cognitive tests in a single parallel process LGC model (**Supplementary Table S3**). There was a significant, negative mean slope for all tests (P < 0.001 except WTAR (P < 0.05)), with the exception of NART where the slope was non-significant. Standard deviation (SD) change per year was calculated for each cognitive test score and ranked in order of most change (1) to least change (13). The four individual processing speed tests showed the largest SD declines over time (range, −0.120 to −0.072), followed by the three visuospatial tests (range, −0.055 to −0.038), the three memory tests (range −0.038 to −0.027), and the three verbal ability tests (range, −0.010 to 0.0001) which showed the least decline. SD change in NART scores was marginally positive but not significantly different from zero (SD change/yr = 0.0001). Model fit indices for Table 3 are shown in **Supplementary Table S4**, alongside those for Tables 4 and 5.

**Table 3.** Latent growth curve models: unstandardized means and variances for the intercept and slope of each cognitive domain, and by APOEe4 carrier status (slopes refer to change from age 70 to age 82)

| Cognitive domain | Intercepts |  | Slopes |  | SD change in each domain |  |
| --- | --- | --- | --- | --- | --- | --- |
|  | Mean (SE) | Variance (SE) | Mean (SE) | Variance (SE) | SD change/yr | Rank order of SD change |
| *All participants* |  |  |  |  |  |  |
| Visuospatial | 15.888 (0.759)*** | 13.711 (1.021) | -0.201 (0.059)** | 0.015 (0.006) | -0.054 | 2 |
| Processing speed | 97.982 (6.013)*** | 21.971 (1.479) | -0.413 (0.046)*** | 0.084 (0.012) | -0.088 | 1 |
| Memory | 72.889 (0.534)*** | 171.046 (17.209) | -0.361 (0.066)*** | 1.722 (0.167) | -0.028 | 3 |
| Verbal ability | 46.757 (1.090)*** | 59.634 (3.009) | -0.022 (0.010)* | 0.016 (0.004) | -0.003 | 4 |
| *APOEε4 non-carriers* |  |  |  |  |  |  |
| Visuospatial | 16.265 (1.053)*** | 13.879 (1.308) | -0.125 (0.048)** | 0.008 (0.005) | -0.033 | 2 |
| Processing speed | 102.548 (7.618)*** | 21.758 (1.760) | -0.318 (0.050)*** | 0.046 (0.010) | -0.068 | 1 |
| Memory | 73.074 (0.653)*** | 182.660 (20.679) | -0.135 (0.072)<sup>NS</sup> | 1.235 (0.155) | -0.010 | 3 |
| Verbal ability | 46.607 (1.373)* | 58.104 (3.691) | -0.026 (0.017)<sup>NS</sup> | 0.019 (0.005) | -0.004 | 4 |
| *APOEε4 carriers* |  |  |  |  |  |  |
| Visuospatial | 14.657 (1.216)*** | 12.850 (1.739) | -0.232 (0.081)** | 0.022 (0.011) | -0.065 | 3 |
| Processing speed | 93.361 (12.045)*** | 22.652 (2.981) | -0.504 (0.084)*** | 0.167 (0.036) | -0.106 | 1 |
| Memory | 71.918 (1.033)*** | 160.519 (35.897) | -0.918 (0.148)*** | 2.714 (0.463) | -0.072 | 2 |
| Verbal ability | 46.416 (2.036)*** | 63.869 (6.017) | -0.021 (0.015)<sup>NS</sup> | 0.015 (0.007) | -0.003 | 4 |

Note: SE, standard error.
Models were run separately for each domain.
Path weights for calculation of the slope factor: Baseline=0; to w2=2.98; to w3=6.75; to w4=9.81; to w5=12.53
SD change/yr is the slope mean divided by the intercept standard deviation; rank order of SD change is from highest (1=most change) to lowest (13=least change)
Model fit statistics are given in Supplementary Table S4
\*p < 0.05; \*\* p < 0.01; \*\*\*p < 0.001

**Table 4.** Latent growth curve models: predictors of intercepts (age 70) and slopes of change (age 70 to 82) where predictors are entered simultaneously (standardised coefficients (Estimate), standard errors (SE), and P-values)

|  | Visuospatial ability |  | Processing speed |  | Memory |  | Verbal ability |  | General cognitive function |  |
| --- | --- | --- | --- | --- | --- | --- | --- | --- | --- | --- |
| Predictors | Estimate (SE) | P values | Estimate (SE) | P values | Estimate (SE) | P values | Estimate (SE) | P values | Estimate (SE) | P values |
| Intercept on |  |  |  |  |  |  |  |  |  |  |
| Age<sup>-</sup> | -0.110 (0.027) | <0.001 | -0.149 (0.027) | <0.001 | -0.157 (0.030) | <0.001 | -0.089 (0.020) | <0.001 | -0.140 (0.021) | <0.001 |
| Sex | -0.261 (0.029) | <0.001 | -0.022 (0.031) | 0.47 | 0.121 (0.033) | <0.001 | 0.002 (0.022) | 0.92 | -0.042 (0.023) | 0.07 |
| Age 11 IQ<sup>+</sup> | 0.494 (0.028) | <0.001 | 0.442 (0.029) | <0.001 | 0.561 (0.033) | <0.001 | 0.566 (0.020) | <0.001 | 0.668 (0.020) | <0.001 |
| Education<sup>+</sup> | 0.109 (0.032) | 0.001 | 0.031 (0.033) | 0.35 | 0.157 (0.036) | <0.001 | 0.239 (0.023) | <0.001 | 0.197 (0.025) | <0.001 |
| Adult SES<sup>-</sup> | -0.124 (0.032) | <0.001 | -0.137 (0.032) | <0.001 | 0.015 (0.036) | 0.69 | -0.110 (0.024) | <0.001 | -0.120 (0.025) | <0.001 |
| Lives alone<sup>-</sup> | 0.029 (0.028) | 0.30 | -0.003 (0.028) | 0.91 | 0.021 (0.031) | 0.50 | -0.033 (0.021) | 0.11 | -0.007 (0.021) | 0.74 |
| Smoking category<sup>-</sup> | -0.065 (0.028) | 0.02 | -0.095 (0.028) | 0.001 | 0.008 (0.031) | 0.80 | -0.032 (0.021) | 0.12 | -0.026 (0.022) | 0.24 |
| Physical activity<sup>+</sup> | 0.039 (0.030) | 0.20 | 0.082 (0.031) | 0.009 | 0.044 (0.034) | 0.20 | -0.009 (0.023) | 0.40 | 0.035 (0.024) | 0.14 |
| Body mass index<sup>-</sup> | 0.084 (0.028) | 0.003 | 0.051 (0.031) | 0.08 | 0.066 (0.032) | 0.03 | -0.053 (0.021) | 0.01 | 0.015 (0.022) | 0.50 |
| Alcohol units, week<sup>+</sup> | 0.000 (0.029) | 0.98 | -0.015 (0.047) | 0.74 | 0.036 (0.032) | 0.26 | -0.019 (0.021) | 0.37 | -0.011 (0.022) | 0.61 |
| APOEε4<sup>-</sup> | -0.100 (0.028) | <0.001 | -0.103 (0.028) | <0.001 | -0.038 (0.031) | 0.23 | 0.001 (0.021) | 0.96 | -0.056 (0.022) | 0.009 |
| Depressive symptoms<sup>-</sup> | -0.059 (0.028) | 0.03 | -0.101 (0.028) | <0.001 | -0.072 (0.031) | 0.02 | -0.018 (0.021) | 0.38 | -0.066 (0.022) | 0.002 |
| CVD<sup>-</sup> | -0.034 (0.028) | 0.22 | -0.069 (0.028) | 0.013 | 0.043 (0.031) | 0.17 | 0.013 (0.020) | 0.52 | -0.005 (0.021) | 0.80 |
| Diabetes<sup>-</sup> | -0.057 (0.028) | 0.04 | -0.057 (0.028) | 0.04 | -0.005 (0.031) | 0.88 | -0.053 (0.021) | 0.01 | -0.055 (0.021) | 0.01 |
| Stroke<sup>-</sup> | -0.024 (0.028) | 0.39 | -0.071 (0.028) | 0.011 | 0.047 (0.031) | 0.12 | 0.028 (0.020) | 0.18 | 0.002 (0.021) | 0.93 |
| Slope on |  |  |  |  |  |  |  |  |  |  |
| Age<sup>-</sup> | 0.111 (0.062) | 0.08 | 0.029 (0.054) | 0.59 | -0.005 (0.044) | 0.91 | 0.262 (0.069) | <0.001 | 0.039 (0.041) | 0.34 |
| Sex | 0.028 (0.067) | 0.68 | 0.075 (0.050) | 0.13 | 0.037 (0.048) | 0.44 | 0.083 (0.066) | 0.21 | 0.040 (0.044) | 0.37 |
| Age 11 IQ<sup>+</sup> | -0.272 (0.077) | <0.001 | -0.044 (0.057) | 0.44 | -0.027 (0.050) | 0.59 | 0.111 (0.070) | 0.11 | -0.062 (0.046) | 0.18 |
| Education<sup>+</sup> | -0.094 (0.072) | 0.19 | 0.011 (0.058) | 0.85 | -0.027 (0.050) | 0.59 | -0.161 (0.073) | 0.03 | -0.057 (0.047) | 0.23 |
| Adult SES<sup>-</sup> | -0.092 (0.072) | 0.20 | 0.025 (0.059) | 0.67 | -0.043 (0.051) | 0.40 | -0.020 (0.069) | 0.77 | -0.010 (0.047) | 0.83 |
| Lives alone<sup>-</sup> | -0.119 (0.065) | 0.07 | -0.037 (0.046) | 0.43 | -0.051 (0.045) | 0.26 | 0.014 (0.062) | 0.83 | -0.031 (0.042) | 0.45 |
| Smoking category<sup>-</sup> | -0.125 (0.072) | 0.08 | 0.022 (0.049) | 0.65 | 0.030 (0.049) | 0.53 | -0.192 (0.070) | 0.007 | -0.021 (0.044) | 0.63 |
| Physical activity<sup>+</sup> | 0.047 (0.068) | 0.49 | 0.020 (0.055) | 0.76 | 0.006 (0.050) | 0.91 | 0.049 (0.069) | 0.47 | 0.062 (0.046) | 0.17 |
| Body mass index<sup>-</sup> | -0.092 (0.067) | 0.17 | -0.073 (0.055) | 0.18 | -0.021 (0.047) | 0.66 | -0.004 (0.064) | 0.95 | -0.036 (0.042) | 0.39 |
| Alcohol units, week<sup>+</sup> | -0.146 (0.074) | 0.048 | 0.019 (0.057) | 0.74 | -0.047 (0.050) | 0.35 | 0.046 (0.071) | 0.51 | -0.015 (0.045) | 0.73 |
| APOEε4<sup>-</sup> | -0.170 (0.065) | 0.009 | -0.211 (0.047) | <0.001 | -0.234 (0.044) | <0.001 | -0.058 (0.061) | 0.35 | -0.246 (0.039) | <0.001 |
| Depressive symptoms<sup>-</sup> | -0.100 (0.065) | 0.12 | -0.060 (0.055) | 0.27 | 0.013 (0.046) | 0.78 | -0.096 (0.063) | 0.13 | -0.071 (0.042) | 0.09 |
| CVD<sup>-</sup> | -0.064 (0.064) | 0.32 | -0.048 (0.046) | 0.30 | 0.005 (0.046) | 0.90 | -0.060 (0.063) | 0.34 | -0.053 (0.042) | 0.21 |
| Diabetes<sup>-</sup> | -0.012 (0.072) | 0.87 | -0.040 (0.050) | 0.42 | -0.088 (0.050) | 0.08 | -0.008 (0.071) | 0.91 | -0.043 (0.044) | 0.33 |
| Stroke<sup>-</sup> | 0.012 (0.074) | 0.87 | 0.083 (0.051) | 0.10 | 0.027 (0.051) | 0.60 | -0.071 (0.071) | 0.32 | 0.039 (0.045) | 0.40 |

Note: SE, standard error; SES, socio-economic status; CVD, cardiovascular disease
<sup>+</sup> hypothesized to have a positive association with cognitive function; <sup>-</sup> hypothesized to have a negative association with cognitive function.
Model estimates are fully standardised.
Path weights for calculation of the slope factor: Baseline=0; to w2=2.98; to w3=6.75; to w4=9.81; to w5=12.53
Models were run separately for each domain; general cognitive function is based on the intercepts and slopes of the four cognitive domains
Boldtype indicates statistical significance following FDR (false discovery rate) correction

#### Latent cognitive domains

Second, we tested whether there was significant ageing-related mean change in each of the four latent cognitive domains for all participants, and then separately by *APOE* e4 carrier status in LGC models (**Table 3**). In the full sample, there was a significant, negative mean slope of ageing-related change across all four cognitive domains. A latent variable of processing speed showed the greatest SD decline per year between age 70 and 82 (SD change/yr = −0.088), followed by visuospatial ability (SD change/yr = −0.054), memory (SD change/yr = −0.028), and verbal ability (SD change/yr = −0.003).

In the *APOE* e4 non-carriers sub-group, the slopes, indicating negative mean change over time, were significant for processing speed (SD change/yr = −0.068) and visuospatial ability (SD change/yr = −0.033) only, but there was little (and non-significant) change in memory (−0.010) or verbal ability (−0.004). In the *APOE* e4 carrier sub-group, the mean slopes were negative and significant for all but verbal ability. Compared to the *APOE* e4 negative group, *APOE* e4 carriers showed greater SD decline in processing speed (SD change/yr = −0.106 vs −0.068), visuospatial ability (SD change/yr = −0.065 vs −0.033), and memory (SD change/yr = −0.072 vs −0.010). The difference was most marked in the slope for memory; *APOE* e4 carriers showed a 7-fold greater SD decline per year compared with *APOE* e4 non-carriers (and in the non-carrier group the slope for memory is non-significant). In contrast with the full sample and the *APOE* e4 non-carriers, memory decline was steeper than visuospatial ability decline in the *APOE* e4-positive group. **Figure 3** presents horizontal bar plots illustrating the SD change/yr in each cognitive test for all participants, and in each cognitive domain for all participants, *APOE* e4 carriers, and *APOE* e4 non-carriers. Formal tests of intercept and slope differences for *APOE* e4 carriers and *APOE* e4 non-carriers are carried out below.

**Figure 3.**
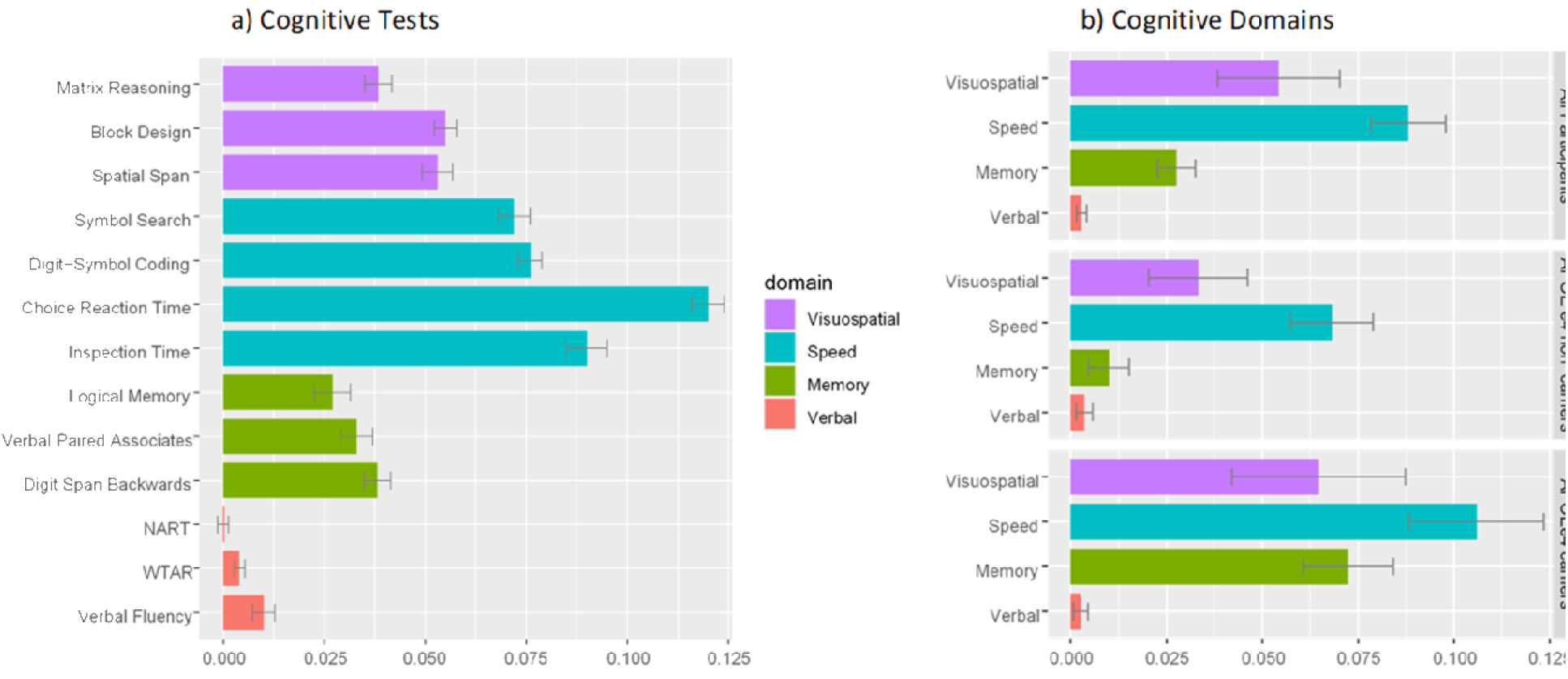
Standard deviation (SD) change per year in a) each cognitive test (grouped by cognitive domain), and b) each cognitive domain (grouped by all participants, and by APOE e4 non-carriers and carriers). SD change per year was derived from latent growth curve models, by calculating the slope mean divided by the intercept SD. SD change per year was converted to +ve values for illustrative purposes, with the exception of NART (National Adult Reading Test) which became –ve. Error bars represent the standard error of SD change per year.

### Predictors of cognitive level and slope

#### Univariate predictors of cognitive level and slope

First, we performed univariate analyses which regressed the intercepts and slopes at the level of each cognitive domain, and then general cognitive function, on all of the predictor variables individually. These univariate (partially-adjusted) models are distinct from the later models featuring multiple risk factors (fully-adjusted) which are the main models of interest. In the univariate models for cognitive ability level at age 70, all of the predictors except living alone were significantly associated with scores on at least one cognitive domain (full results are shown in **Supplementary Table S5)**. In the univariate models for cognitive slope, only *APOE* e4 status, alcohol, smoking, and age 11 IQ were significant predictors of decline across selected domains. *APOE* e4 carriers were more likely to show decline between age 70 and age 82 in visuospatial ability (β = −0.185, P = 0.005), speed (β = −0.215, P < 0.001), memory (β = −0.235, P < 0.001), and general cognitive ability (β = −0.233, P < 0.001). Smoking was associated with more decline in verbal ability (β = −0.203, P = 0.004) only, and a higher alcohol intake was associated with more decline in visuospatial ability only (β = −0.183, P = 0.015). Finally, a higher childhood cognitive ability (β = -0.252, P = 0.001) was associated with more decline in visuospatial ability only.

#### Multivariate predictors of cognitive level at age 70

Next, we ran multivariate models to simultaneously estimate the effect of multiple risk factors on cognitive level at age 70. When all fifteen predictors were modelled at the same time, thirteen (not living alone or alcohol intake) made a significant contribution to the variability in cognitive ability level at age 70 (i.e. the intercept) in at least one of the cognitive domains (upper section, **Table 4**). Performance on all four cognitive domains and the general factor of cognitive function was associated with age (within-wave differences) (range, standardised beta (β) = −0.089 to −0.157, P < 0.001) and age 11 IQ (range β = 0.442 to 0.668, P < 0.001); age 11 IQ accounted for the most variance in cognitive level of any of the predictors, with the largest effect size (β = 0.668) for general cognitive function. Education and SES predicted performance in the general factor, and three out of four of the domains (no association between education-speed and between SES-memory), with an average (β) effect size across the four domains of -0.176 and -0.123, respectively. The directions of effects were as expected, such that individuals with better age 70 cognitive ability level were younger, had a higher childhood intelligence, were more educated, and were from more professional occupational classes. Male sex (β = 0.261, P < 0.001) was a predictor of better visuospatial ability level, and female sex was a predictor of better memory level (β = 0.121, P < 0.001), but sex was not a significant predictor of general cognitive function.

Healthy lifestyle factors were selectively associated with better cognitive ability at age 70: more physical activity (β = 0.082, P = 0.009) and less smoking (β = −0.095, P = −0.001) with better processing speed. A higher BMI (a measure of obesity) was associated with a lower verbal ability (β = −0.053, P = 0.01) but conversely with higher visuospatial ability (β = 0.084, P = 0.003). Alcohol intake did not significantly predict age 70 cognitive ability in any domain. None of the lifestyle factors measured were significantly associated with general cognitive function in the multivariate model. *APOE* e4 positive carrier status predicted poorer visuospatial ability (β = −0.100, P < 0.001), processing speed (β = −0.103, P < 0.001) and general cognitive function (β = −0.056, P = 0.009) at age 70. History of disease was associated with lower cognitive scores but not consistently across domains: CVD (β = −0.069, P = 0.013) and stroke (β = −0.071, P = 0.011), were associated with lower processing speed, in addition to a non-FDR-significant association with diabetes (β = −0.057, P = 0.04). Diabetes was associated with lower verbal ability (β = −0.053, P = 0.01) and general cognitive function (β = −-0.055, P = 0.01). Depressive symptoms were associated with lower processing speed (β = −0.101, P < 0.001) and general cognitive function (β = −0.066, P = 0.002). Notably, many of the previous univariate associations between individual predictors and cognitive level at age 70 (across selected domains) became non-significant in the multivariate models.

#### Multivariate predictors of cognitive slope between age 70 and 82

In contrast to cognitive level at age 70, we found that few predictors were associated with longitudinal cognitive change between age 70 and 82 (as shown in **Table 4** for slope, lower section) once all fifteen predictors were entered simultaneously. *APOE* e4 carrier status accounted for the most variability in cognitive slopes. Possessing the *APOE* e4 allele was associated with significantly steeper decline in visuospatial ability (β = −0.170, P = 0.009), processing speed (β = −0.211, P < 0.001), memory (β = −0.234, P < 0.001), and general cognitive function (β = −0.246, P < 0.001), but not with verbal ability (β = −0.058, P = 0.35). Moreover, *APOE* e4 was the *only unique* significant predictor of cognitive change in processing speed, memory, and general cognitive function, with resultant effect sizes markedly larger in magnitude than any of the other variables. Other than being an *APOE* e4 allele carrier, a steeper slope in visuospatial ability was also associated with a having a higher age 11 IQ (β = −0.272, P < 0.001). The only predictors of a steeper verbal ability slope were more smoking (β = −0.192, P = 0.007), and contrary to expectations, a lower age (β = 0.262, P < 0.001). Comparisons between the univariate and multivariate predictor models for cognitive slope indicate that the univariate association between higher alcohol intake and greater decline in visuospatial ability (β = −0.183, P = 0.015) was non-significant in the multivariate model (β = −0.146, P = 0.05).

**Figure 4** illustrates the unique variance (R^2^) accounted for by the fifteen predictor variables in **Table 4** for each cognitive domain, versus a matched set of simulated random variables. These comparisons allow us to check whether our predictor group performed better than the same number of null variables, and are presented as stacked barplots showing the real data (in colour) and random data (in grey). The overall R^2^ for the set of real predictors was significantly larger than the null scenario across the domains: visuospatial ability (real = 20%, null = 4%); processing speed (real 8%= null = 2%); memory (real = 8%, null = 1%); verbal ability (real = 16%, null = 4%); general cognitive function (real = 9%, null = 2%).

**Figure 4.**
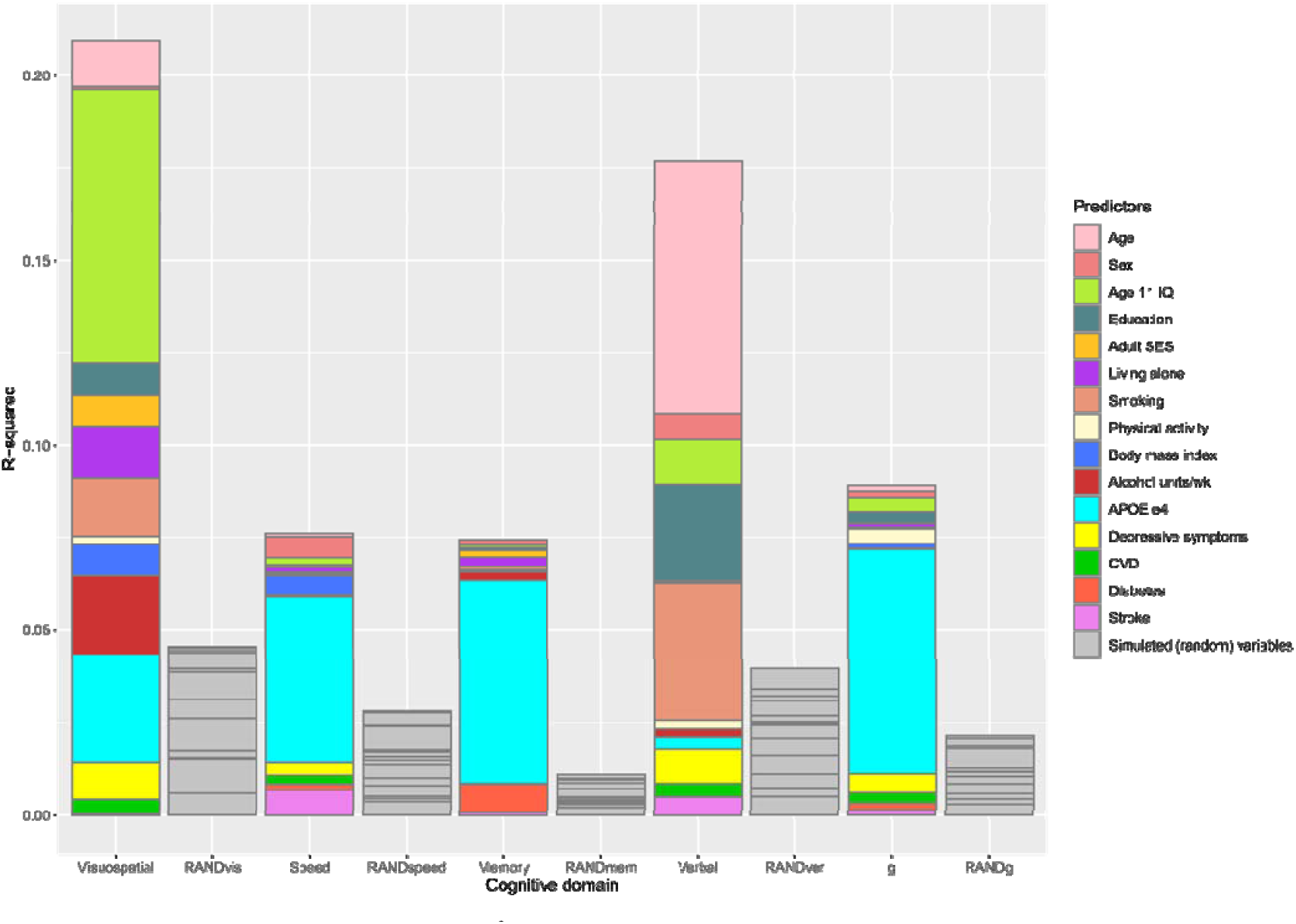
Stacked barplots showing the unique variance (R^2^) in cognitive domain slopes explained by the predictor variables in the multivariate models (Table 5). Grey columns show the R^2^ explained by the same number of simulated (random) variables in each cognitive domain as a comparison.

#### Sensitivity analyses

We performed three sensitivity analyses to determine whether our results were driven by: participants who developed dementia by the age 82 assessment (N = 24); low MMSE scorers at one or more testing waves (N = 46); and deaths (N = 403). We found no substantive differences between the results of the sensitivity analyses (reported in **Supplementary Material Tables S6-8**) and those reported above. The only notable result of these exclusions was an attenuation in effect sizes for the *APOE* e4 associations with visuospatial ability slope, of 46%, 22%, and 34%, respectively, across the three analyses, which were no longer significant at P < 0.05.

## Discussion

We characterised longitudinal changes across major domains of cognitive functioning over a 12-year period, modelling a comprehensive set of cognitive tests administered five times at 3-yearly intervals— allowing a robust examination of rates of decline—in a birth cohort of older adults for whom childhood IQ scores are available. Using a multivariate approach, we examined the relative contributions of determinants of individual differences in age 70-cognitive level and age 70 to 82-cognitive change, using fifteen of the most commonly used candidate risk factors in the field of cognitive ageing. Our key finding is that *APOE* e4 status was the single most important factor determining longitudinal cognitive decline when all of the predictors were modelled simultaneously. Carriers of the *APOE* e4 allele show significantly steeper declines across the three ‘fluid’ domains of memory, processing speed, and visuospatial ability, compared to non-carriers, even after adjusting for many other potential predictors which were strong correlates of age 70 cognitive level (including childhood IQ, education, adult socioeconomic status, lifestyle, and health). *APOE* e4 status was the sole predictor of decline in general cognitive function—with a moderate to large effect size of 0.25 [57]—comparable in magnitude, for instance, to the reduction in risk of dying from head injuries associated with wearing a cycling helmet [58]. This contrasts with the relatively modest cross-sectional associations between *APOE* e4 and cognitive functioning at age 70 which suggests that the effect of *APOE* e4 on cognitive deficits becomes more manifest in later life. These findings are striking given that when many other candidate predictors of cognitive ageing slope are entered en masse, their unique contributions account for relatively small proportions of variance, beyond variation in *APOE* e4 status, and might indicate an increasing genetic influence on cognitive outcomes as individuals’ progress into their eighth and ninth decades of life.

The presence of faster rates of decline in *APOE* e4 carriers, across several different domains of cognitive functioning, adds valuable new data to the debate on whether *APOE* e4 influences cognitive ageing. Our findings stand in contrast with some studies which report null findings such as the Australian PATH study [59], and the HALCyon programme which provided only very limited evidence of an effect of *APOE* e4 on a test of word recall, but not on other cognitive measures [19]. Discrepancies in findings may reflect differences in sample age; both samples were considerably younger than the present study, perhaps too young to show e4-related decrements. Our results extend prior work that does find an effect of *APOE* e4 in the following ways. First, we report that *APOE* e4 exerts broad and general adverse effects on cognitive functioning, typically only reported in cross-sectional meta-analytic data across many piecemeal studies [25], but not in a single longitudinal cohort study. Second, we found a particularly deleterious effect of *APOE* e4 on memory decline, consistent with two single-candidate studies using a single memory test [23, 60]. Here, we show this effect is robust to simultaneous adjustments in a multi-candidate study, and reliable across a broad cognitive trait of memory, captured by the latent domain. Third, we show that the relationship between *APOE* e4 and long-term cognitive decline is largely independent of childhood cognitive ability, an important confound (but rarely available measure) in studies of cognitive ageing [61]. Fourth, we were able to show that the *APOE* e4 allele affects age-related cognitive decline independently of possible cognitive impairment, dementia, and deaths to follow up, suggesting that this relationship is present, not just in dementia and Alzheimer’s Disease [17, 62], but in cognitively ‘healthy’ individuals.

Our results suggest that differences in cognitive functioning between e4 and non-e4 carriers become more pronounced with advancing age, regardless of any pathological changes. This finding aligns with earlier reports of an age effect of *APOE* e4 on cognition across the lifespan in single-determinant studies, with associations rarely seen in those <70 years [19, 23]. In 19,594 participants of the Health and Retirement study, age-stratified analyses showed there was relatively no effect of being a carrier at age 50-59, compared to age 80 and above, where there was an almost 2.5 year difference in ‘cognitive age’, a marker of cognitive functioning, compared with non-carriers [63]. Age effects of the *APOE* e4 allele support theories suggesting that the presence of the allele leads to reductions in neural protection and repair, and that carriers are more vulnerable to damage accumulated over their lifetime [64].

We found limited evidence in the LBC1936 that individual health behaviours alter rates of decline between ages 70-82 years when modelled in tandem. Those with a history of smoking showed faster declines in verbal ability, in agreement with a large body of evidence documenting the detrimental effects of smoking on cognition and brain health [27, 29-30], though the change in this crystallised domain was minimal over time. One major question for the field of cognitive ageing is whether various lifestyle choices all compete for a limited opportunity to enhance cognitive function or whether the effects could be additive, as part of a synergistic lifestyle pattern [65-66]. While there were few individual effects, Figure 4 makes it clear that together, lifestyle predictors account for a greater amount of the variance in cognitive decline than might be attributed to chance. In accordance with a ‘marginal gains’ theory of cognitive ageing [28], individual differences in cognitive trajectories among our sample, probably reflect an accumulation of small influences from numerous lifestyle (and other) factors. Though the magnitude of the observed associations between the various individual lifestyle factors and cognitive change were mostly small, if these associations represent a causal effect, their cumulative efforts are likely to have significance for cognitive health at the population level.

Consistent with previous studies [36, 67], a higher childhood IQ—the strongest predictor of higher (cross-sectional) age-70 cognitive level in our sample—did not confer an advantage in terms of protection from steeper declines in the long-term. The only evidence of an effect of early-life cognitive ability was a faster decline in visuospatial ability in those with a higher childhood IQ. This counterintuitive finding was surprising but not unusual, and may indicate regression to the mean, that is, a consequence of higher ability individuals performing relatively more poorly on tests with known ceiling effects when followed longitudinally [68]. Nevertheless, the current study benefits from knowing individuals’ cognitive starting point. Early-life cognition is associated with a subsequent cascade of social, behavioural and clinical effects [69] such that children with higher cognitive ability tend to become brighter and healthier adults [28]. Being able to remove this confound from our models is important to reduce the likelihood of the observed associations being artefacts of the relationship between childhood IQ and healthy life markers. In doing so, our findings help to address an important issue in cognitive ageing research, namely, distinguishing differential preservation from preserved differentiation [8, 70]. With the clear exception of *APOE*, our results support the preserved differentiation of cognitive function only —whereby *level* of ability is a manifestation of prior ability— but not differential preservation (which leads to differences in subsequent rates of decline).

Declines in processing speed between age 70 and 82 were greater than those of the other domains which supports the theory that processing speed is the core issue responsible for deficits in performance on complex cognitive measures in ageing populations [71-73]. Memory declined less steeply, across the whole sample, than processing speed and visuospatial ability, even in the ninth decade when one might expect to see more pronounced changes in this domain [74]. However, memory tests repeated longitudinally are subject to practice effects, such that participants may be able to improve or maintain their tests scores in spite of a cognitive decline [75]. Despite the potential of practice effects to obscure the variance in memory performance measured over time (e.g. in tests containing memorable information in stories or word lists), ageing effects were still present in the data, and if anything, they may lead to an underestimation of true effect sizes. Verbal ability showed evidence of stability with age, as expected [76-78]. Nevertheless, the observation of concomitant rises in word knowledge alongside marked declines in other cognitive measures with age, is still of empirical value.

### Strengths and limitations

The major strength of the LBC1936 is an unusually comprehensive cognitive battery, enabling good characterisation of cognitive domains across later life, and the ability to account for childhood cognitive ability, which is uncommon in studies of cognitive ageing. Identical tests and testing location were used at five sampling occasions over a 12 year follow-up period, covering an age-critical window in later life for accelerated cognitive decline. The multivariate design of the study addresses the multicollinearity of a range of life-course predictors. Modelling latent cognitive variables reduced the influence of potential measurement error inherent in using single cognitive tests. We further improved the robustness of our results by using FDR-adjustment for multiple associations, thereby reducing the chance of type I errors., and conducting sensitivity tests for incident dementia and death.

The study results should be interpreted with several limitations in mind. As with any longitudinal study, a key limitation was survival bias such that those who remained in the study were healthier older individuals with more education, a higher SES, and a lower prevalence of comorbidities. However, the modest 20% attrition rate over each successive follow-up is comparable to those of other highly valuable longitudinal cohort studies with repeated assessments, such as the Swedish National Study on Aging [79] and the English Longitudinal Study of Ageing [80]. Using FIML in our LGC analyses partly addresses the issue of attrition by including all available data from each time-point, not just completers, resulting in less biased estimates. Some physiological processes preceding cognitive decline may occur earlier in life, therefore, mid-life measures of some risk factors, e.g. physical activity and BMI, may be more important at predicting rapid cognitive decline than measures obtained later in life [81-83], but are not available in this cohort. We were also unable to further explore associations according to *APOE* e4 allele variations, given that the numbers of individuals in each allele group were insufficient to conduct further comparisons between e2, e3 and e4 genotypes. We acknowledge that our cognitive intercept at age 70 is likely to be a conflation of both intercept and some degree of slope (i.e. cognitive ageing experienced up to that point). Without knowing individuals’ mid-age (reflecting *peak* cognitive function) to older-age trajectories, we cannot fully address the issue of preserved differentiation vs differed preservation, though childhood IQ functions as a good proxy measure given its stability across the lifespan [84]. Finally, as a volunteer sample, the LBC1936 represent a well-educated, generally healthy group, which might preclude the generalisation of these findings to the broader ageing population, and as such, replication in other larger samples is warranted.

In summary, we found that *APOE* e4 status was the single most important predictor of longitudinal cognitive decline from age 70 to 82, when fifteen potential predictors were modelled simultaneously, despite there being many life-course correlates of cognitive level at age 70. *APOE* e4 allele carriers experienced significantly steeper 12-year declines across the three ‘fluid’ domains of memory, processing speed, and visuospatial ability, and a general factor of cognitive function, than non-carriers, denoting an increasingly widening gap in cognitive functioning as individuals’ progress into older age. Our findings suggest that (1) when many other candidate predictors of cognitive ageing slope are entered en masse, their unique contributions account for relatively small proportions of variance, beyond variation in *APOE* e4 carrier status, (2) *APOE* e4 status is important for identifying those a greater risk for accelerated cognitive ageing, even among ostensibly healthy individuals.

## Data Availability

LBC1936 data is available upon reasonable request (https://www.ed.ac.uk/lothian-birth-cohorts/data-access-collaboration)

## Acknowledgements

This research was funded in whole, or in part, by the Wellcome Trust [221890/Z/20/Z]. For the purpose of open access, the author has applied a CC BY public copyright licence to any Author Accepted Manuscript version arising from this submission.

We gratefully acknowledge the contributions of the LBC1936 participants and members of the LBC1936 research team who collect and manage the LBC data. We also thank the Genetics Core staff at the Edinburgh Clinical Research Facility. The LBC1936 is supported by Age UK [The Disconnected Mind], the Medical Research Council [G0701120, G1001245, MR/M01311/1, MR/R024065/1] and the University of Edinburgh. SRC and IJD were additionally supported by a National Institutes of Health (NIH) research grant R01AG054628, and IJD was also supported by the Dementias Platform UK [MR/L015382/1].SRC is supported by a Sir Henry Dale Fellowship jointly funded by the Wellcome Trust and the Royal Society (Grant Number 221890/Z/20/Z).

## Box 1

Advantages of the study design

1. Unusually comprehensive cognitive battery with several high-quality tests for each cognitive domain.
2. 12 year follow-up—5 testing periods—using identical tests, equipment, and testing location.
3. Cognitive testing across an important period from age 70, when cognitive ageing becomes pertinent, to age 82, when risk of rapid decline and dementia dramatically increases.
4. Record of general cognitive ability at age 11.
5. Multiple (correlated) candidate determinants are included in mutually-adjusted models enabling estimates of relative contributions of each predictor to cognitive change.
6. It is well-powered.
7. There are sensitivity tests for incident dementia and death.

## Footnotes

1 To test for non-linear components of cognitive ageing, separate measurement models included a quadratic term with quadratic increasing slope weightings. Visual inspection of the mean trajectory of memory test scores from age 70-82 indicated that memory might best be modelled using a non-linear factor of change (to account for the rise in mean test scores in the initial waves of testing, followed by a fall towards the end of the follow-up). However, these models did not converge and are not discussed further.

